# A comprehensive assessment of statin discontinuation among patients who concurrently initiate statins and CYP3A4-inhibitor drugs; a multistate transition model

**DOI:** 10.1101/2021.05.13.21252626

**Authors:** Macarius M. Donneyong, Yuxi Zhu, Teng-Jen Chang, Pengyue Zhang, Yiting Li, Katherine M. Hunold, ChienWei Chiang, Kathleen Unroe, Jeffrey M. Caterino, Lang Li

## Abstract

**Aims:** To describe the 1-year direct and indirect transition probabilities to premature discontinuation of statin therapy after concurrently initiating statins and CYP3A4-inhibitor drugs.

**Methods:** A retrospective new-user cohort study design was used to identify (N=160828) patients who concurrently initiated CYP3A4-inhibitors (diltiazem, ketoconazole, clarithromycin, others) and CYP3A4-metabolized statins (statin DDI exposed, n = 104774) vs. other statins (unexposed, n = 56054) from the MarketScan Commercial claims database (2012 – 2017). These groups were matched (2:1) through propensity score-matching techniques. We applied a multistate transition model to compare the 1-year transition probabilities involving four distinct states (start, adverse drug events [ADEs], discontinuation of CYP3A4-inhibitor drugs, and discontinuation of statin therapy) between those exposed to statin DDIs, vs. unexposed. Statistically significant differences were assessed by comparing the 95% confidence intervals (CIs) of probabilities.

**Results:** Patients exposed to statin DDIs, vs. unexposed, were significantly less likely to discontinue statin therapy (71.4 [95% CI: 71.1, 71.6] vs. 73.3 [95% CI: 72.9, 73.6]) but more likely to experience an ADE (3.4 [95% CI: 3.3, 3.5] vs. 3.2 [95% CI: 3.1, 3.3]) and discontinue with CYP3A4-inhibitor therapy (21.0 [95% CI: 20.8, 21.3] vs. 19.5 [95% CI: 19.2, 19.8]) directly after concurrently starting stains and CYP3A. Subsequent to experiencing an ADE, those exposed to statin DDIs were still less likely to discontinue statin therapy but were significantly more likely to discontinue CYP3A4-inhibitor therapy.

**Conclusion:** While statin DDI exposure was associated with higher likelihood of ADEs, this did not increase the risk of premature statin discontinuation among patients exposed to statin DDIs, versus unexposed.

## Background

Statins are the most frequently used medications for controlling cholesterol levels and hence are able to reduce the risk of several cardiovascular diseases (CVD). About 25% - 53% of patients discontinue statin therapy within a year after initiation [1-4]. Discontinuation of statin therapy is a major risk factor of cardiovascular diseases, stroke and death among patients who need statin therapy [5-9]. Statin-induced adverse drug events (ADEs), perceived or actual, are one of the major reasons for discontinuation of statin therapy [10].

Polypharmacy is common among patients with indication for statin therapy that exposes patients to ADEs, especially rhabdomyolysis and myalgia, due to drug-drug interactions (DDIs). Atorvastatin, lovastatin and simvastatin are metabolized by the Cytochrome P450 3A4 (CYP3A4) enzymes. Pharmacokinetic data shows that systemic exposure to statins is elevated when these CYP3A4-metabolized statins are taken concurrently with CYP3A4-inhibitor drugs. Higher systemic exposure to statins could potentially increase the risk of statin-induced ADEs and subsequently lead to premature discontinuation of statin therapy. However, some data from real-world populations suggests that the effects of pharmacokinetic statin DDIs may not translate to higher risk of statin-induced ADEs. The effects of drug interactions between CYP3A4-metabolized statins and individual CYP3A4-inhibitors on premature statin discontinuation has not been well investigated. This data is important for helping clinicians select statin-CYP3A4-inhibitor drug pairs.

The primary objective of this study was to describe the 1-year probabilities of the potential states (start, ADEs, discontinuation of CYP3A4-inhibitor drugs, and discontinuation of statin therapy) that patients may transition to after concurrently initiating statins and CYP3A4-inhibitor drugs before finally discontinuing statin therapy. We were also interested in assessing for the presence of potential differences in transition probabilities among subgroup of patients who used specific individual CYP3A4-inhibitors. We hypothesized that the probability of premature statin discontinuation would be higher among patients who concurrently initiated CYP3A4-inibitor drugs and CYP3A4-metabolized statins (statin DDI exposed) compared to those who concurrently initiated other statins that are not metabolized by CYP3A4 enzymes (unexposed). We also hypothesized that the statin DDI exposed group would have a higher likelihood of experiencing ADEs which would lead to higher probabilities of statin discontinuation in this group compared to the unexposed statin user group.

## METHODS

### Cohort selection

A retrospective cohort study design was used to identify statin users from the IBM^®^ MarketScan^®^ claims database. We had approval to use this data through a Data User Agreement between The Ohio State University (OSU) and IBM. Further, The OSU IRB did not constitute the project to require human subjects review. To minimize confounding by indication we restricted the analysis to only new statin initiators by requiring users to have had no records of prior fills of any statins in the 183 days period leading to the eligible statin fill date (index date) for cohort entry (**Figure 1a/1b**) [11, 12]. For this reason, we included only participants with at least 183 days of continuous enrollment (i.e. no loss in health coverage of >30 days) in a health plan. Patients whose index statin fill had <30 days’ supply were also excluded to ensure that they were concurrently exposed to statins and CYP3A-inhibiting drugs. Among eligible statin users we excluded those <18 years old on the statin index date. Further, the primary analysis were restricted to only patients who were continuously enrolled during the 365 days after concurrently using statins and CYP3A4-inhibitors.

**Figure 1a:**
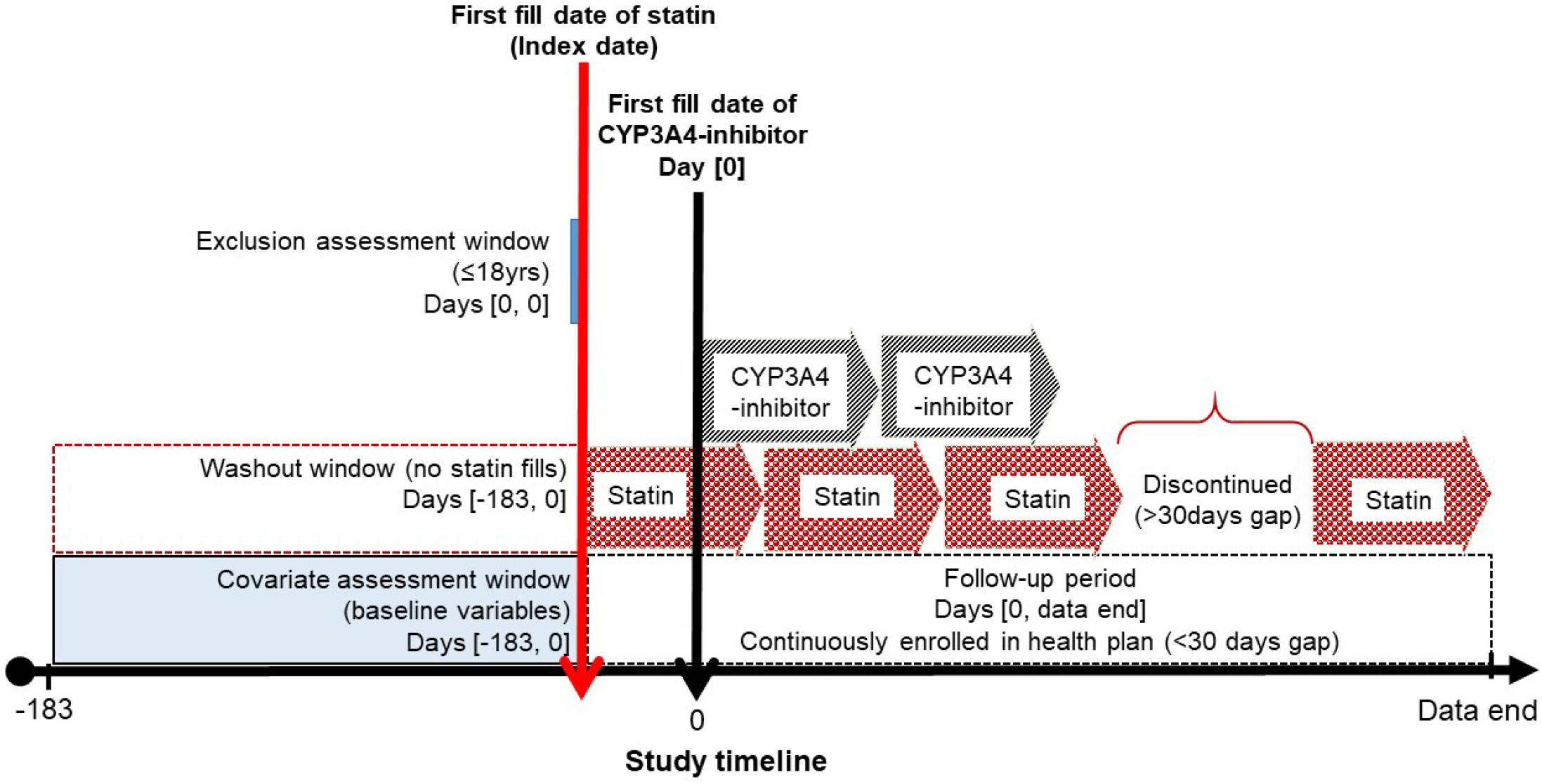
New users of statins who subsequently initiated a CYP3A4-inhibitor

**Figure 1b:**
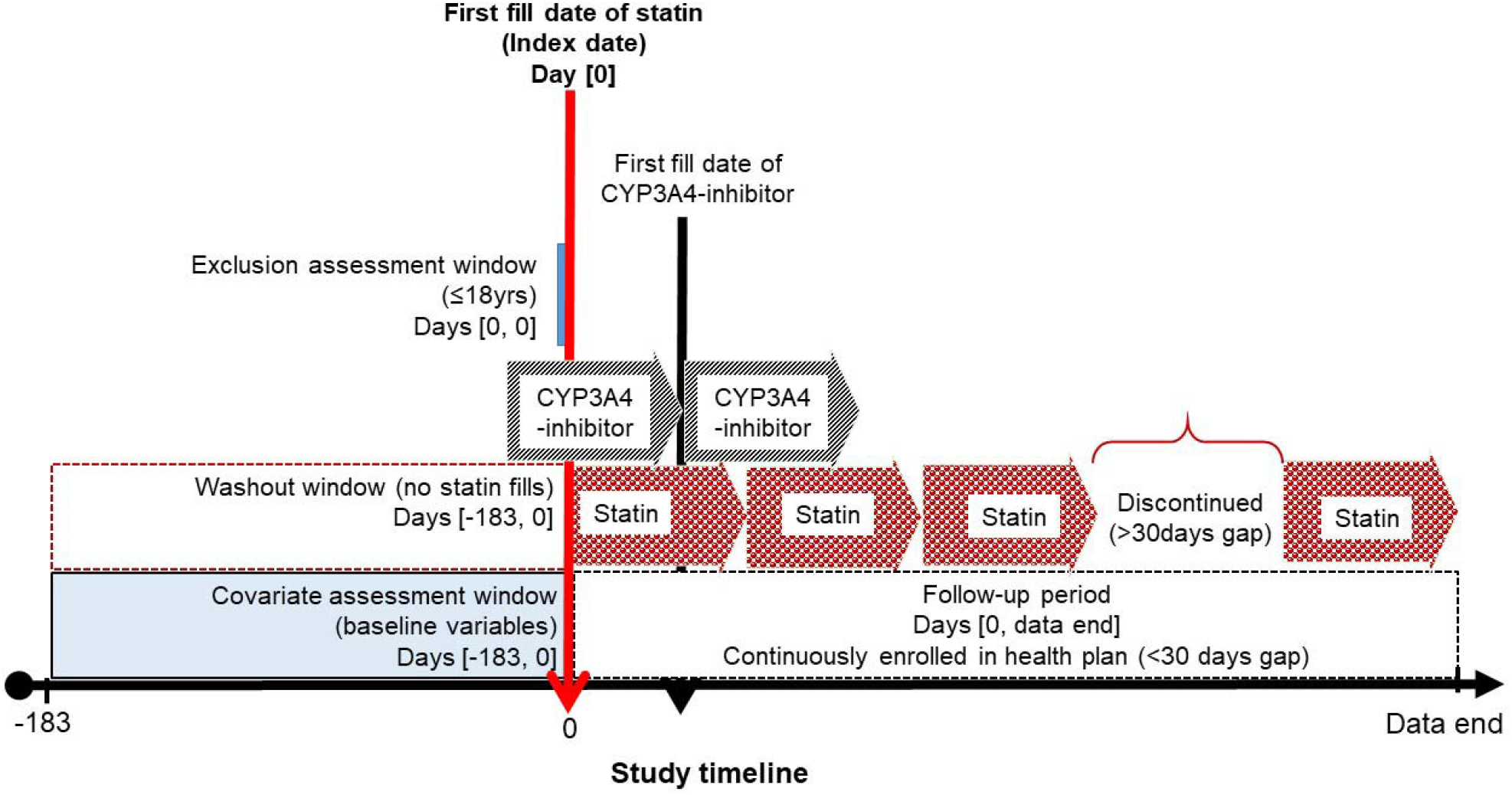
New users of statins who were already using a CYP3A4-inhibitor

### Definition of concomitant exposure to statins and CYP3CA-inhibitor drugs

A recent meta-analysis by Nguyen et. al. (2018) reported that the concomitant use of statins and the following CYP3A4-inhibitor drugs resulted in statin-induced adverse events including rhabdomyolysis and myalgia: clarithromycin, erythromycin, nefazodone and diltiazem [13]. For patients who filled any of these CYP3A-inhibitors after statin initiation date (index date), concurrent use was defined as an overlap in the days’ supply of the index statin with the fill date of the CYP3A4-inhibitor drug (**Figure 1a**). Conversely, if the statin index date occurred after the CYP3A4-inhibitor fill date, then concurrent use was defined as an overlap in the days’ supply of the inhibitor drug with the statin index date (Cohort 2) (**Figure 1b**). Among the derived cohorts of concurrent users of statins and CYP3A-inhibitor drug initiators, we considered patients as DDI exposed if they concurrently initiated CYP3A4-metabolized statins (atorvastatin, lovastatin, simvastatin), those who concurrently used other statins (fluvastatin, pitavastatin, pravastatin, rosuvastatin) and CYP3A-inhibitor drugs were considered as unexposed to DDIs. The unexposed statin users served as an active comparator group since there is no known drug-drug interactions between these statins and CYP3A4-inhibitor drugs. The active user comparator study design is a powerful strategy for controlling for confounding by indication in observational study design [14]. Both cohorts 1 and 2 were combined to implement the statistical analysis described in later sections.

### Follow up

Follow up for outcomes started on the fill date of a concomitant CYP3A-inhibitor drug during the index statin exposure (based days’ supply) in Cohort 1 (**Figure 1a**) through to the end of data availability. On the otherhand, outcomes were assessed beginning on the date that patients initiated a new statin in Cohort 2 (**Figure 1b**) through to the end of data availability.

### Outcomes

Premature statin discontinuation: Statin discontinuation within 365 days after initiation was considered as premature discontinuation of statin therapy. We defined statin discontinuation as a gap of ≥90 days between the end of days’ supply of the last filled prescription and a subsequent statin fill or end of data availability, whichever comes first.

#### Intermediary outcomes

Statin-induced ADEs: Statin-induced ADEs were defined using a previously validated algorithm for identifying statin intolerance from claims data and included: musculoskeletal (rhabdomyolysis and myopathy), gastrointestinal-related ADEs, cognitive dysfunction, other [15].

Switching: Switching between statins was defined as the filling of a different statin from the opposite exposure group after concomitant exposure, e.g. from starting with a CYP3A4-metabolized statin (atorvastatin, lovastatin or simvastatin) before filling a different statin (fluvastatin, pitavastatin, pravastatin or rosuvastatin), or vice versa.

Discontinuation of CYP3A4-inhibitor drug therapy: this was defined as a gap of ≥90 days between the end of days’ supply of the last filled CYP3A4-inhibitor prescription and a subsequent fill or end of data availability, whichever comes first. Although we use the term “discontinuation”, a discontinuation of use of clarithromycin and erythromycin may actually be an indication of the completion of therapy. However, for ease of readability we use the term discontinuation of CYP3A4-inhibitors to refer to both discontinuation and completion of therapy.

### Covariates

Several demographic, clinical, characteristics of index statins and inhibitor drugs (dose, days’ supply) use of other chronic conditions medications), Charleson-Elixhauser combined comorbidity index and healthcare utilization factors were measured at baseline (**Table 2**). These covariates were measured during the 183 day period prior to the start of concurrent exposure to new statins and CYP3A4-inhibitors (**Figure 1a/1b**).

### Multistate transition structure

Figure 2. is a graphical representation of the multistate transition state model comprising of four states: start (of concurrent exposure to statins and CYP3A4-inhibitors), ADEs, discontinuation of CYP3A4-inhibitor drugs, and discontinuation of statin therapy. This generated six different transitions (from→to): 1) start→ADEs; 2) start→ discontinuation of CYP3A4-inhibitor; 3) start→discontinuation of statin therapy; 4) ADE→ discontinuation of CYP3A4-inhibitor; 5) ADE→ discontinuation of statin therapy; and 6) discontinuation of CYP3A4-inhibitor→ discontinuation of statin therapy. In sensitivity analysis we introduced switching between CYP3A4-metabolized statins and other statins as a competing risk for discontinuation of statin therapy, i.e. rather than discontinue with statins, clinicians might switch patients to a different statin if there are any clinical concerns.

### Statistical analysis

We applied propensity score matching techniques to control for potential confounding effects of measured covariates listed in **Table 1**. Because there were nearly twice as many statin DDI exposed as there were unexposed patients, we matched these groups in a 2:1 ratio. We assessed the balance of covariates between the statin DDI exposed and unexposed groups with standardized difference test at the start of concurrent exposure to statins and CYP3A4-inhibitors. We further assessed the balance of covariates between the exposure groups during each transition period given that the sample and distributions of covariates vary over transition periods. The goal of the statistical analysis was to generate the transition probabilities for the overall population and among the statin DDI exposed and unexposed subgroups. First, we described the distribution of baseline covariates between the statin DDI exposed and unexposed groups during each transition. Second, a Markov model was used to calculate the 1-year transition probabilities among the propensity score matched sample [16]. The Markov model assumes that future states depend on the history only through the present state; e.g. for the start→ADE→statin discontinuation path in our specified multistate transition model, the probability of statin discontinuation is dependent on the occurrence of ADE. The overall and statin DDI exposure group-specific transition probabilities and their associated standard errors were generated through the *mstate* package in R [16]. We then constructed 95% confidence intervals for each transition probability using the associated standard errors. We assessed statistical significance between in the difference between transition probabilities for the statin DDI exposed and unexposed groups by comparing the 95% CIs for the respective probabilities. A non-overlap in the 95% CIs between any two probabilities was deemed statistically significant. Because we were interested in assessing differences in the impact of interactions between statins and specific individual CYP3A4-inhibitors, we repeated the analysis among subgroups who concurrently used specific individual CYP3A4-inhibitors and statins.

**Table 1:** Distribution of baseline covariates between statin users exposed to statin drug-drug interactions versus not exposed.

| Baseline covariates | Frequency (%) / mean (SD) |  | Standardized difference |
| --- | --- | --- | --- |
|  | Statin DDI Unexposed (N = 56054) | Statin DDI Exposed (N = 104774) |  |
| **Demographics** |  |  |  |
| Age groups (years) |  |  | 0.03 |
| <45 | 3673 ( 6.6) | 7129 ( 6.8) |  |
| 45 - 54 | 10204 (18.2) | 18737 (17.9) |  |
| 55 - 64 | 21045 (37.5) | 38247 (36.5) |  |
| 65 - 74 | 10989 (19.6) | 20601 (19.7) |  |
| 75 and above | 10143 (18.1) | 20060 (19.1) |  |
| Sex |  |  |  |
| Female | 26906 (0.50) | 52387 (0.50) | 0.05 |
| Calendar year of index statin |  |  | 0.05 |
| 2012 | 9754 (17.4) | 17265 (16.5) |  |
| 2013 | 16036 (28.6) | 28459 (27.2) |  |
| 2014 | 13683 (24.4) | 26145 (25.0) |  |
| 2015 | 9151 (16.3) | 17865 (17.1) |  |
| 2016 | 7430 (13.3) | 15040 (14.4) |  |
| **Index statin-related factors** |  |  |  |
| Days' supply of index statin (days) |  |  | 0.02 |
| <60 | 25969 (46.3) | 47885 (45.7) |  |
| 60 - 90 | 513 ( 0.9) | 845 ( 0.8) |  |
| >90 | 29572 (52.8) | 56044 (53.5) |  |
| Dose of index statin (mg) |  |  | 0.23 |
| <10 | 7134 (12.7) | 6770 ( 6.5) |  |
| 10 - 19 | 12438 (22.2) | 26738 (25.5) |  |
| 20 - 40 | 17361 (31.0) | 36678 (35.0) |  |
| >40 | 19121 (34.1) | 34588 (33.0) |  |
| **Index CYP3A4-inhibitor factors** |  |  |  |
| Index CYP3A4-inhibitors concurrently initiated with statins<sup>a</sup> |  |  | 0.13 |
| Clarithromycin | 4561 ( 8.1) | 8465 ( 8.1) |  |
| Diltiazem Hydrochloride | 38386 (68.5) | 71766 (68.5) |  |
| Ketoconazole | 8904 (15.9) | 19519 (18.6) |  |
| others | 4203 ( 7.5) | 5024 ( 4.8) |  |
| Duration of episode of CYP3A4-inhibitor treatment (days) |  |  | 0.04 |
| <30 | 9005 (16.1) | 17730 (16.9) |  |
| 30 - 89 | 10254 (18.3) | 19957 (19.0) |  |
| 90 - 269 | 18896 (33.7) | 34857 (33.3) |  |
| >270 | 17899 (31.9) | 32230 (30.8) |  |
| **Disease conditions that make up statin-induced ADEs** |  |  |  |
| Cognitive impairment | 171 ( 0.3) | 329 ( 0.3) | 0.00 |
| Gastrointestinal bleeding | 6776 (12.1) | 12362 (11.8) | 0.01 |
| Musculoskeletal disease | 11522 (20.6) | 20982 (20.0) | 0.01 |
| Myopathy | 1749 ( 3.1) | 3165 ( 3.0) | 0.01 |
| Others | 1936 ( 3.5) | 3404 ( 3.2) | 0.01 |
| **Chronic disease conditions** |  |  |  |
| Alzheimer's disease | 296 ( 0.5) | 623 ( 0.6) | 0.01 |
| Asthma | 8421 (15.0) | 15593 (14.9) | 0.00 |
| Cancer | 7128 (12.7) | 13307 (12.7) | 0.00 |
| Coronary artery disease | 10612 (18.9) | 19414 (18.5) | 0.01 |
| Depression | 3616 ( 6.5) | 6685 ( 6.4) | 0.00 |
| Diabetes | 10508 (18.7) | 19393 (18.5) | 0.01 |
| Hypertension | 36080 (64.4) | 66857 (63.8) | 0.01 |
| Kidney diseases | 1583 ( 2.8) | 3119 ( 3.0) | 0.01 |
| Liver diseases | 1771 ( 3.2) | 3165 ( 3.0) | 0.01 |
| Myocardial infarction | 533 ( 1.0) | 1081 ( 1.0) | 0.01 |
| Peripheral heart disease | 550 ( 1.0) | 1101 ( 1.1) | 0.01 |
| Stroke | 340 ( 0.6) | 727 ( 0.7) | 0.01 |
| Valvular heart disease | 133 ( 0.2) | 264 ( 0.3) | 0.00 |
| Combined comorbidity score (mean (SD)) | 18956 (33.8) | 36916 (35.2) | 0.03 |
| **Chronic disease medications** |  |  |  |
| Angiotensin receptive blockers | 8103 (14.5) | 14593 (13.9) | 0.02 |
| Angiotensin converting enzyme inhibitors | 12265 (21.9) | 23907 (22.8) | 0.02 |
| Anticoagulants | 8001 (14.3) | 15835 (15.1) | 0.02 |
| Antiplatelets | 4346 ( 7.8) | 8202 ( 7.8) | 0.00 |
| Antipsychotics | 1092 ( 1.9) | 2046 ( 2.0) | 0.00 |
| Benzodiazepines | 8743 (15.6) | 15669 (15.0) | 0.02 |
| Beta blockers | 16273 (29.0) | 31042 (29.6) | 0.01 |
| Bisphosphonates | 1732 ( 3.1) | 3176 ( 3.0) | 0.00 |
| Calcium channel blockers | 39125 (69.8) | 72233 (68.9) | 0.02 |
| Diuretics | 21790 (38.9) | 40517 (38.7) | 0.00 |
| **Healthcare utilization** |  |  |  |
| Counts of unique drugs prescribed (mean (SD)) | 8.63 (5.30) | 8.32 (5.19) | 0.06 |
| Counts of unique physician visits (mean (SD)) | 1.06 (0.24) | 1.06 (0.24) | 0.01 |
| Counts of hospital days (mean (SD)) | 1.55 (4.21) | 1.54 (3.88) | 0.00 |
| Counts of unique pharmacies (mean (SD)) | 1.26 (0.85) | 1.26 (0.81) | 0.01 |
| Paid copay/coinsurance associated for index office visit | 17029 (30.4) | 30925 (29.5) | 0.02 |

Abbreviation: SD, standard deviation
SD ≤0.1 implies a balance between the statin DDI exposed and unexposed groups
<sup>a</sup>Not included in the propensity-score matching

**Figure 2:**
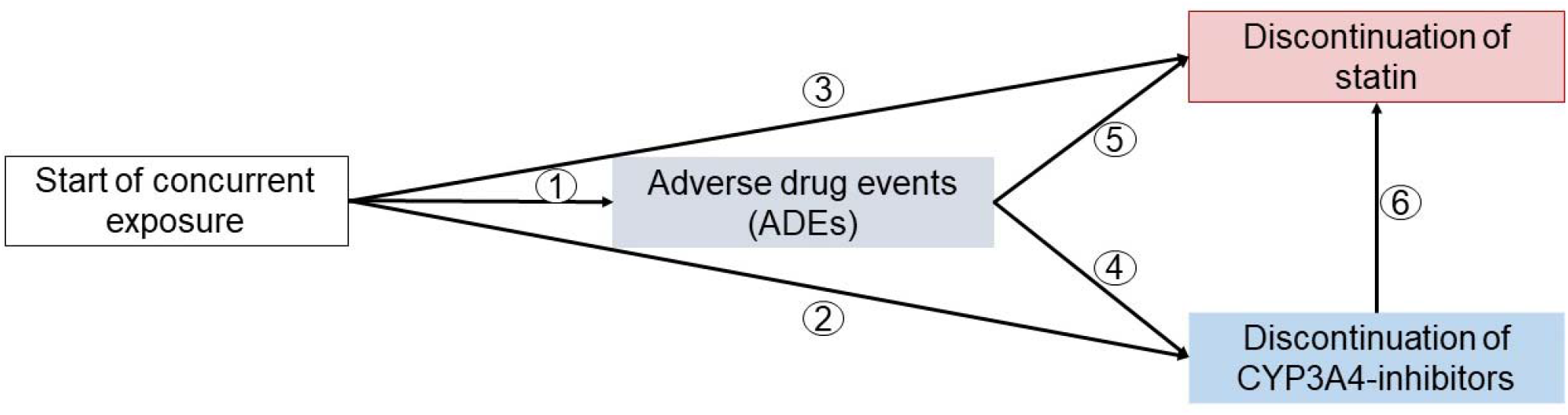
A graphical representation of four-state multistate transition state model for assessing events after concurrent exposure to statins and CYP3A4-inhibitors.

## Results

We identified 160828 patients who concurrently initiated CYP3CA-inhibitors and CYP3CA-metabolized statins (n = 104774) vs. other statins (n = 56054). Over two-thirds (68.5%) of statin users in both the statin DDI exposed and unexposed groups concurrently initiated diltiazem; the rest initiated ketoconazole (18.6% exposed; 15.9% unexposed), clarithromycin (8.1% exposed; 8.1% unexposed) and other inhibitors (4.8% exposed; 7.5% unexposed) (**Table 1**). All measured covariates, except the dosage of the index statin, were balanced (standardized difference <0.1) after matching, before the start of concurrent exposure and during each of the six transition periods. The statin DDI exposed group initiated relatively lower dosages (<10mg) of statins compared to the unexposed group; 6.5% vs. 12.7%.

### Transition probabilities, overall and by subgroups of CYP3A4-inhibitors

**Table 2** is a summary of the 1-year transition probabilities, overall and stratified by individual CYP3A4-inhibitors. It was feasible to only produce stratified results for diltiazem hydrochloride, ketoconazole and clarithromycin which were concurrently initiated by over 90% of statin users. The overall probability of transitioning from start of concurrent exposure to statin discontinuation was 72.0%; higher probabilities were observed for those who concurrently initiated statins and ketoconazole (74.6%) or clarithromycin (77.2%). The transition probabilities from start of exposure to ADEs were very low, 3.3%. However, concurrent users of statins and diltiazem were more likely (4.1%) to experience ADEs compared to statin users who concurrently initiated ketoconazole (0.8%) or clarithromycin (0.3%). The concurrent use of statins and ketoconazole (24.5%) or clarithromycin (22.4%) were associated with relatively higher probabilities of discontinuation of CYP3A4-inhibitor therapy, compared to the concurrent use of diltiazem (19.8%). This was expected given that both ketoconazole and clarithromycin usually have a short course of treatment. The probabilities of transitioning to statin discontinuation increased after the occurrence of ADEs, compared to from start of exposure: overall (75.6%), diltiazem (74.1%), ketoconazole (76.6%), clarithromycin (78.2%). However, patients were relatively less likely to transition to a state of discontinuation of CYP3A4-inhibitor therapies after the occurrence of ADEs: overall (17.3%), diltiazem (17.3%), ketoconazole (20.2%), clarithromycin (20.1%). The probabilities to transition to statin discontinuation were further increased after patients discontinued with CYP3A4-inhibitor therapies with the highest probabilities observed among those who concurrently initiated diltiazem (82.3%). The inclusion of switching between statins as a competing event for statin discontinuation did not change these results in a meaningful way (**Supplemental Table 1**).

**Table 2:** One-year transition probabilities of statin-induced adverse drug events, discontinuation of CYP3A4-inhibitors and discontinuation of statin therapy, overall and by subgroups of CYP3A4-inhibitors.

| Transition |  | One-year transition probabilities, % (95% CI) |  |  |  |
| --- | --- | --- | --- | --- | --- |
| From | To | Overall | Diltiazem | Ketoconazole | Clarithromycin |
| Start | Adverse drug event | 3.3 (3.2, 3.4) | 4.1 (4.0, 4.2) | 0.8 (0.7, 0.9) | 0.3 (0.2, 0.4) |
| Start | CYP3A4-inhibitor discontinuation | 20.5 (20.3, 20.7) | 19.8 (19.5, 20.0) | 24.5 (23.9, 25.0) | 22.5 (21.7, 23.3) |
| Start | Statin discontinuation | 72.0 (71.8, 72.2) | 70.4 (70.1, 70.6) | 74.6 (74.1, 75.2) | 77.2 (76.4, 77.9) |
| Adverse drug event | CYP3A4-inhibitor discontinuation | 17.3 (17.0, 17.6) | 17.3 (17.0, 17.7) | 20.2 (19.5, 20.8) | 20.1 (19.3, 21.0) |
| Adverse drug event | Statin discontinuation | 75.6 (75.3, 75.9) | 74.1 (73.6, 74.5) | 76.6 (76.0, 77.2) | 78.2 (77.4, 79.0) |
| CYP3A4-inhibitor discontinuation | Statin discontinuation | 78.8 (78.5, 79.1) | 82.3 (81.8, 82.8) | 75.7 (75.1, 76.3) | 76.3 (75.5, 77.2) |

### Comparing the transition probabilities between statin DDI exposed vs. unexposed groups

We observed several small but statistically significant differences in the transition probabilities between the stain DDI exposed and unexposed groups. The transition probabilities between the statin DDI exposed and unexposed groups are visually represented in **Figure 3. Table 3** contain these probabilities together with 95% CIs, for the overall population and by subgroups individual CYP3A4-inhibitors. We observed small but statistically significant differences in transition probabilities between patients exposed to statin DDIs (concurrent use of CYP3A4-metabolized statins and CYP3A4-inhibitors) vs. those who were unexposed (concurrent use of statins not metabolized by CYP3A4 enzymes and CYP3A4-inhibitors). After concurrently initiating statins and CYP3A4-inhibitors, those exposed to statin DDIs, vs. unexposed, were significantly less likely to discontinue statin therapy (71.4 [95% CI: 71.1, 71.6] vs. 73.3 [95% CI: 72.9, 73.6]) but more likely to experience an ADE (3.4 [95% CI: 3.3, 3.5] vs. 3.2 [95% CI: 3.1, 3.3]) and discontinue with CYP3A4-inhibitor therapy (21.0 [95% CI: 20.8, 21.3] vs. 19.5 [95% CI: 19.2, 19.8]). Similar differences were observed among subgroups who concurrently initiated statins and only diltiazem or only ketoconazole but not among those who concurrently initiated clarithromycin. Subsequent to experiencing an ADE, those exposed to statin DDIs were still less likely to discontinue statin therapy (74.9 [95% CI: 74.5, 75.4] vs. 76.8 [95% CI: 76.2, 77.4]) but were significantly more likely to discontinue CYP3A4-inhibitor therapy (17.7 [95% CI: 17.4, 18.0] vs. 16.6 [95% CI: 16.1, 17.0]). Again, similar comparative differences were observed among diltiazem and ketoconazole user but not among clarithromycin users. Patients who were exposed to statin DDIs were also less likely to discontinue statin therapy after they had discontinued with CYP3A4-inhibitor therapy: 78.3 (95% CI:78.0, 78.7) vs. 79.7 (95% CI: 79.1, 80.2) among the unexposed group, overall and among diltiazem and ketoconazole users subgroups but not among clarithromycin users.

**Table 3:** Comparison of the one-year transition probabilities between statin users exposed to statin drug-drug interactions versus not exposed, overall and by subgroups of CYP3A4-inhibitors.

| Transition |  | One-year transition probabilities, % (95% CI) |  |
| --- | --- | --- | --- |
| From | To | Statin DDI exposed | Statin DDI unexposed |
| Overall (statins plus any CYP3A4-inhibitors) |  |  |  |
| Start | Adverse drug event | 3.4 (3.3, 3.5) | 3.2 (3.1, 3.3) |
| Start | CYP3A4-inhibitor discontinuation | 21.0 (20.8, 21.3) | 19.5 (19.2, 19.8) |
| Start | Statin discontinuation | 71.4 (71.1, 71.6) | 73.3 (72.9, 73.6) |
| Adverse drug event | CYP3A4-inhibitor discontinuation | 17.7 (17.4, 18.0) | 16.6 (16.1, 17.0) |
| Adverse drug event | Statin discontinuation | 74.9 (74.5, 75.4) | 76.8 (76.2, 77.4) |
| CYP3A4-inhibitor discontinuation | Statin discontinuation | 78.3 (78.0, 78.7) | 79.7 (79.1, 80.2) |
| **Diltiazem (statins plus only diltiazem)** |  |  |  |
| Start | Adverse drug event | 4.2 (4.0, 4.3) | 3.9 (3.7, 4.1) |
| Start | CYP3A4-inhibitor discontinuation | 20.4 (20.1, 20.7) | 18.6 (18.2, 19.0) |
| Start | Statin discontinuation | 69.5 (69.1, 69.8) | 72.0 (71.6, 72.5) |
| Adverse drug event | CYP3A4-inhibitor discontinuation | 17.9 (17.4, 18.3) | 16.3 (15.7, 16.8) |
| Adverse drug event | Statin discontinuation | 73.1 (72.6, 73.7) | 75.8 (75.0, 76.5) |
| CYP3A4-inhibitor discontinuation | Statin discontinuation | 81.8 (81.1, 82.4) | 83.3 (82.6, 84.1) |
| **Ketoconazole (statins plus only ketoconazole)** |  |  |  |
| Start | Adverse drug event | 0.8 (0.7, 0.9) | 0.7 (0.6, 0.8) |
| Start | CYP3A4-inhibitor discontinuation | 25.2 (24.5, 25.8) | 23.0 (22.0, 23.9) |
| Start | Statin discontinuation | 73.9 (73.3, 74.6) | 76.2 (75.3, 77.1) |
| Adverse drug event | CYP3A4-inhibitor discontinuation | 20.8 (20.1, 21.6) | 18.7 (17.6, 19.8) |
| Adverse drug event | Statin discontinuation | 75.7 (74.9, 76.4) | 78.5 (77.3, 79.6) |
| CYP3A4-inhibitor discontinuation | Statin discontinuation | 74.9 (74.2, 75.6) | 77.4 (76.3, 78.5) |
| **Clarithromycin (statins plus only clarithromycin)** |  |  |  |
| Start | Adverse drug event | 0.3 (0.2, 0.4) | 0.3 (0.2, 0.3) |
| Start | CYP3A4-inhibitor discontinuation | 22.4 (21.5, 23.4) | 22.6 (21.4, 23.9) |
| Start | Statin discontinuation | 77.3 (76.3, 78.2) | 77.0 (75.7, 78.3) |
| Adverse drug event | CYP3A4-inhibitor discontinuation | 19.8 (18.8, 20.8) | 20.8 (19.4, 22.2) |
| Adverse drug event | Statin discontinuation | 78.5 (77.5, 79.5) | 77.7 (76.4, 79.1) |
| CYP3A4-inhibitor discontinuation | Statin discontinuation | 76.6 (75.6, 77.6) | 75.9 (74.5, 77.3) |

**Figure 3:**
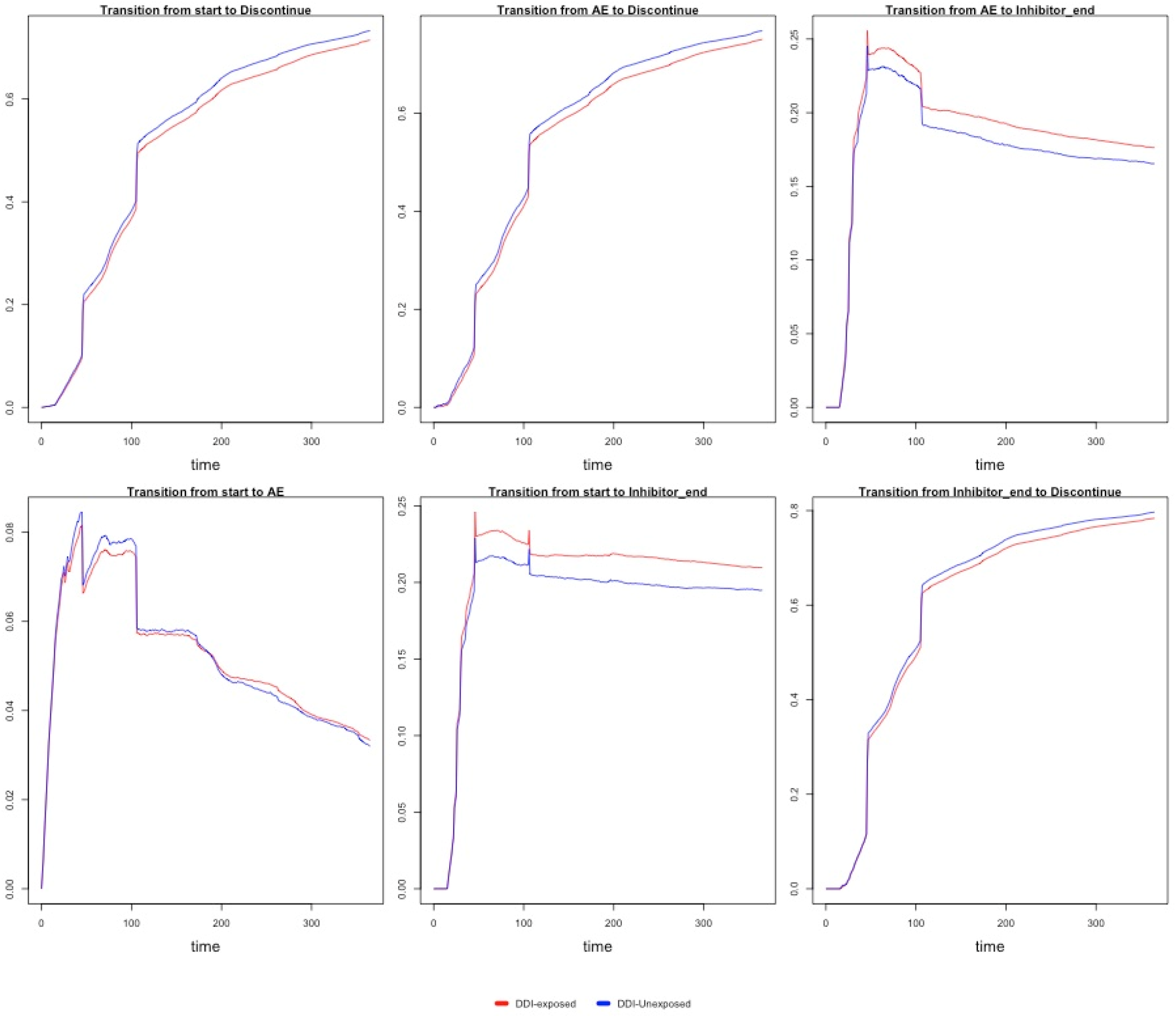
Comparison of the one-year transition probabilities between statin users exposed to statin drug-drug interactions versus not exposed.

## Discussion

Our data confirms that, overall, patients are at an increased risk of prematurely discontinuing statin therapy within a year after concurrently initiating any statin and a CYP3A4-inhibitor. Further, we have confirmed that the occurrence of ADEs was associated with an additional increased probability of premature discontinuation of statin therapy overall. However, contrary to our hypothesis, concurrent use of CYP3A4-metabolized statins and CYP3A4-inhibitors was rather associated with slightly lower probability of statin discontinuation both directly after exposure and subsequent to ADEs. Interestingly, patients rarely switched from CYP3A4-metabolized statins to other statins even after the occurrence of statin-induced ADEs. We have shown that diltiazem and ketoconazole are the top two most frequently used CYP3A4-inhibitors that drive the observed probabilities of statin discontinuation and ADEs after exposure to statin DDIs.

There are no published data on the risk of premature discontinuation of statin therapy among statin users who concurrently initiate CYP3A4-inhibitor drugs. A recent study among older adults in Australia reported that 64% of statin users discontinued statin therapy after one year of initiation [17]. However, several other previously published data reported much lower 1-year prevalence of statin discontinuation, 13% - 35% among patients using [18]. In the context of these lower prevalence of statin discontinuation, the 72.0% probability observed in our data confirms that the concurrent initiation of statins and CYP3A4-inhibitor drugs is a major risk factor of statin discontinuation, irrespective of whether patients are using CYP3A4-metabolized statins or other statins [19, 20].

Although the probability of premature discontinuation was only slightly lower among the statin DDI exposed group, compared to the unexposed group, this finding was contrary to our stated hypothesis. This unexpected finding could partially be explained by the relatively higher rate of discontinuation of CYP3A4-inhibitor drug therapy among the statin DDI exposed group, compared to the unexposed comparator group. It is plausible that clinicians preemptively withdrew patients from CYP3A4-inhibitor drug therapy after discovering that they may be at risk for ADEs due to the concomitant use of CYP3A4-metabolized statins and CYP3A4-inhibitor drugs. This theory is supported by our finding of higher probabilities of discontinuation of CYP3A4-inhibitor drug therapies among the statin DDI exposed group, compared to the unexposed group, both directly after concurrent initiation and subsequent to the occurrence of ADEs. It appears that clinicians preferred to request patients to discontinue CYP3A4-inhbitor therapy rather than statin therapy, especially when patients were concurrently exposed to CYP3A4-metabolized statins and CYP3A4-inhibitor drugs. The lack of difference in the probability of switching from a CYP3A4-metabolized statin to those not metabolized by these enzymes after the occurrence ADEs suggests that discontinuation of CYP3A4-inhibitors is a potential effective strategy for reducing the risk of premature statin discontinuation among patients exposed to statin DDIs.

The role of the dose of the index statin on statin discontinuation warranted further exploration especially given that the statin DDI exposed group initiated relatively lower doses (<10mg) of statins compared to the unexposed group. We compared the probability of statin discontinuation between the statin DDI exposed vs. unexposed groups across levels of statin doses after start and after ADEs. The probability of statin discontinuation after start was significantly lower among the statin DDI exposed, vs. unexposed groups, only at lower statin doses (<10mg); 69.1% (95% CI: 68.0%, 70.2%) vs. 76.7% (95% CI: 75.7%, 77.7%). This difference narrowed across increasing levels of statin doses and became null at the highest dose level (>40mg); 72.0% (95% CI: 71.5%, 72.5%) vs. 71.8% (95% CI: 71.1%, 72.4%). Similar differences in the probabilities of statin discontinuation between the exposed and unexposed were observed after the occurrence of ADEs at doses <10mg (72.8% [95% CI: 71.2%, 74.4%] vs. 80.4% [95% CI: 79.0%, 81.8%]) compared to >40mg (75.7% [95% CI: 75.0%, 76.5%] vs. 75.4% [95% CI: 74.4%, 76.4%]). This data suggests that patients exposed to statin DDIs were only less likely to discontinue statin therapy, compared to unexposed, only at low doses of statins. If confirmed this data is confirmed in other studies, initiating patients at low statin doses might help prevent premature statin discontinuation among patients exposed to statin DDIs.

### Clinical implications

Our findings imply that patients who concurrently use statins and CYP3A4-inhibitor drugs are at higher risk of premature discontinuation of statins irrespective of whether they are concurrently using CYP3A4-metabolized statins or not. This group of patients should be targeted as a high-risk group for interventions to reduce premature discontinuation of statin therapy. Because the probability of statin discontinuation was high even without an antecedent ADE event, clinicians should consider the implementation of interventions as soon as a patient is identified to be concurrently using statins and CYP3A4-inhibitor drugs. Based on our data, clinicians and patients could consider short-term use of CYP3A4-inhibitor drugs, especially diltiazem and ketoconazole. The discontinuation of diltiazem and ketoconazole resulted in relatively lower risk of statin discontinuation among patients exposed to statin DDIs. The risk-benefit effect of short-term use of CYP3A4-inhibitors should, however, be carefully evaluated before adopting this as a strategy to prevent premature statin discontinuation.

### Limitations

Our data should be interpreted with caution because of the following study limitations: 1) There is lack of information on whether discontinuation was intentional or not in the MarketScan claims data, this is true for all administrative claims data. Our analysis assumes that majority of patients discontinued statin therapy intentionally on their own or at the behest of a clinician. If this is true, then our study provides data that can help guide the development of interventions to modify this behavior. Our findings, however, may not be very relevant for patients who discontinue statin therapy unintentionally due to forgetfulness, lack of access due to cost, etc. 2) Because we operationalized statin discontinuation based on refill gaps (30, 60, 90 days), this definition may misclassify patients if they purchased statins out of pocket without using their health insurance. We neither expect this factor to be pervasive nor to be differential between the two statin user groups compared in our analysis. 3) Although we applied propensity score matching techniques to reduce potential confounding by measured baseline covariates, the confounding effects of unmeasured factors, especial SES, could potentially bias the effects estimates observed in the comparator analysis. 4) The MarketScan data comprises of commercially insured populations, therefore our findings may have limited generalizability to uninsured and older adults (65 years and older).

### Strengths

Inspite of the aforementioned limitations our study has several strengths. First, we used a new user active-comparative study design to implement our analysis. This approach is a robust strategy for reducing bias and confounding by indication in pharmacoepidemiological research [14]. Second, the application of propensity score matching techniques enabled us balance baseline covariates between the statin DDI exposed and unexposed groups. Third, the application of a multistate transition model enabled us to generate the most comprehensive data on the direct and indirect probabilities and comparative risks of statin discontinuation. This type of information is critical for optimizing adherence and statin treatment outcomes among patients who have indications for concurrent treatment with statins and CYP3A4-inhibitors. Fourth, we have shown that diltiazem and ketoconazole are frequently used with statins and are associated with higher probabilities of statin discontinuation and statin-induced ADEs. Our results suggest the discontinuation of these two CYP3A4-inhibitors could potential mitigate premature statin discontinuation among those exposed to statin DDIs.

## Conclusion

Overall, concurrent users of statins and CYP3A4-inhibitor drugs, especially diltiazem and ketoconazole, were likely to discontinue statin therapy with or without the occurrence of statin-induced ADEs. However, there was no evidence of higher risk of discontinuation among concurrent users of CYP3A4-inhibitors and CYP3A4-metabolized statins, vs. other statins, before or after the occurrence of ADEs. The discontinuation of CYP3A4-inhibitors (diltiazem and ketoconazole) is a potential strategy to mitigate premature discontinuation of statin therapy if confirmed through intervention studies. Such future intervention studies should seek to better understand the risk-benefit trade-off of discontinuing treatment with CYP3A4-inhibitor drugs as an intervention to prevent premature statin discontinuation.

## Data Availability

The MarketScan claims data can be requested from IBM: https://www.ibm.com/products/marketscan-research-databases

