## Supplemental Tables for "A comprehensive assessment of statin discontinuation among patients who concurrently initiate statins and CYP3A4-inhibitor drugs; a multistate transition model"

**Table 3**: Comparison of the one-year transition probabilities between statin users exposed to statin drug-drug interactions versus not exposed, overall and by subgroups of CYP3A4-inhibitors.

| **Transition** | | **One-year transition probabilities, % (95% CI)** | | |
| --- | --- | --- | --- | --- |
|  |  | **Overall** | **Comparison between** | |
| **From** | **To** |  | **Statin DDI exposed** | **Statin DDI unexposed** |
| **Overall (statins plus any CYP3A4-inhibitors)** |  |  |  |  |
| Start | Statin discontinuation | 70.5 (70.3, 70.8) | 70.1 (69.8, 70.4) | 71.4 (71, 71.7) |
| Start | Adverse drug event | 3.3 (3.2, 3.3) | 3.3 (3.2, 3.4) | 3.2 (3.0, 3.3) |
| Start | Switching between statins | 1.5 (1.5, 1.6) | 1.3 (1.2, 1.4) | 2.0 (1.9, 2.1) |
| Start | CYP3A4-inhibitor discontinuation | 20.5 (20.3, 20.7) | 21.0 (20.8, 21.3) | 19.5 (19.2, 19.8) |
| CYP3A4-inhibitor discontinuation | Statin discontinuation | 78.5 (78.2, 78.8) | 78.1 (77.8, 78.5) | 79.3 (78.7, 79.8) |
| Adverse drug event | Statin discontinuation | 73.4 (72.9, 73.9) | 72.9 (72.2, 73.6) | 74.3 (73.5, 75.2) |
| Adverse drug event | Switching between statins | 2.6 (2.0, 3.2) | 2.5 (1.7, 3.2) | 2.8 (2.0, 3.7) |
| Adverse drug event | CYP3A4-inhibitor discontinuation | 17.1 (16.8, 17.4) | 17.4 (17.1, 17.8) | 16.4 (16, 16.9) |
| **Diltiazem (statins plus only diltiazem)** |  |  |  |  |
| Start | Statin discontinuation | 68.7 (68.4, 69) | 68.1 (67.7, 68.4) | 69.9 (69.4, 70.3) |
| Start | Adverse drug event | 4.0 (3.9, 4.1) | 4.1 (4.0, 4.3) | 3.8 (3.6, 4.0) |
| Start | Switching between statins | 1.8 (1.7, 1.9) | 1.5 (1.4, 1.6) | 2.3 (2.2, 2.5) |
| Start | CYP3A4-inhibitor discontinuation | 19.7 (19.5, 19.9) | 20.3 (20.0, 20.6) | 18.5 (18.1, 18.9) |
| CYP3A4-inhibitor discontinuation | Statin discontinuation | 82.1 (81.6, 82.6) | 81.6 (80.9, 82.2) | 83.0 (82.3, 83.8) |
| Adverse drug event | Statin discontinuation | 71.6 (70.9, 72.3) | 70.8 (69.9, 71.7) | 73.1 (72.1, 74.2) |
| Adverse drug event | Switching between statins | 3.1 (2.3, 3.8) | 3.0 (2.0, 4.0) | 3.2 (2.2, 4.1) |
| Adverse drug event | CYP3A4-inhibitor discontinuation | 16.9 (16.6, 17.3) | 17.5 (17.0, 17.9) | 16.0 (15.4, 16.6) |
| **Ketoconazole (statins plus only ketoconazole)** |  |  |  |  |
| Start | Statin discontinuation | 73.7 (73.2, 74.3) | 73.3 (72.6, 73.9) | 74.8 (73.8, 75.7) |
| Start | Adverse drug event | 0.8 (0.7, 0.8) | 0.8 (0.7, 0.9) | 0.7 (0.6, 0.8) |
| Start | Switching between statins | 0.7 (0.6, 0.8) | 0.5 (0.4, 0.6) | 1.0 (0.8, 1.3) |
| Start | CYP3A4-inhibitor discontinuation | 24.7 (24.1, 25.2) | 25.3 (24.6, 25.9) | 23.4 (22.4, 24.3) |
| CYP3A4-inhibitor discontinuation | Statin discontinuation | 75.4 (74.8, 76.0) | 74.7 (74.0, 75.4) | 76.8 (75.6, 77.9) |
| Adverse drug event | Statin discontinuation | 75.4 (74.7, 76.2) | 75.0 (74.2, 75.8) | 76.4 (74.9, 77.9) |
| Adverse drug event | Switching between statins | 1.1 (0.6, 1.6) | 0.6 (0.2, 1.1) | 2.0 (0.8, 3.1) |
| Adverse drug event | CYP3A4-inhibitor discontinuation | 20.3 (19.7, 20.9) | 20.9 (20.2, 21.7) | 19.0 (17.8, 20.1) |
| **Clarithromycin (statins plus only clarithromycin)** |  |  |  |  |
| Start | Statin discontinuation | 76.3 (75.5, 77.0) | 76.4 (75.5, 77.4) | 76.0 (74.7, 77.3) |
| Start | Adverse drug event | 0.3 (0.2, 0.3) | 0.3 (0.2, 0.4) | 0.3 (0.2, 0.3) |
| Start | Switching between statins | 0.7 (0.6, 0.8) | 0.6 (0.5, 0.8) | 0.8 (0.6, 1.1) |
| Start | CYP3A4-inhibitor discontinuation | 22.7 (22.0, 23.5) | 22.7 (21.7, 23.6) | 22.9 (21.6, 24.1) |
| CYP3A4-inhibitor discontinuation | Statin discontinuation | 76.0 (75.1, 76.8) | 76.2 (75.2, 77.3) | 75.5 (74.1, 76.9) |
| Adverse drug event | Statin discontinuation | 77.1 (76.2, 78.0) | 77.3 (76.2, 78.4) | 76.7 (75.1, 78.2) |
| Adverse drug event | Switching between statins | 1.0 (0.5, 1.5) | 1.0 (0.4, 1.7) | 0.9 (0.0, 1.8) |
| Adverse drug event | CYP3A4-inhibitor discontinuation | 20.3 (19.5, 21.1) | 19.9 (18.9, 21.0) | 21.0 (19.6, 22.4) |
